# Modeling Indirect Protection from Typhoid Conjugate Vaccines in the TyVAC-Bangladesh Trial

**DOI:** 10.64898/2025.12.05.25341721

**Authors:** Jo Walker, Farhana Khanam, Xinxue Liu, Merryn Voysey, Md. Asif Ahsan, Md. Golam Firoj, Nazmul Hasan Rajib, Prasanta Kumar Biswas, Andrew J. Pollard, K. Zaman, Firdausi Qadri, John D. Clemens, Virginia E. Pitzer

## Abstract

In the TyVAC-Bangladesh cluster-randomized trial, a single dose of typhoid conjugate vaccine (TCV) in children aged 9 months to <16 years old conferred strong total protection against typhoid fever. However, indirect protection among unvaccinated cluster residents, including older age groups ineligible for vaccination, was modest and did not reach statistical significance. This result raises questions about the effectiveness of TCV against transmission of *Salmonella* Typhi and whether TCV introduction in endemic settings will induce herd effects. We hypothesize that the low indirect protection observed in the TyVAC-Bangladesh study could be explained by some combination of transmission across cluster boundaries (contamination), limited direct vaccine protection against infection, and insufficient population-wide vaccine coverage. To evaluate these hypotheses, we simulated the trial using two transmission dynamic models: a primary model fit exclusively to clinical surveillance data from the Bangladesh study site, and an alternative model simultaneously fit to clinical and serological data. Vaccinating 64% of eligible children (the coverage level achieved in the TyVAC-Bangladesh trial) induced 0% to 67% indirect protection in the primary model, and 0% to 54% indirect protection in the alternative model, depending on the transmissibility of chronic carriers and TCV efficacy against S. Typhi infection. The results of the TyVAC-Bangladesh trial were consistent with a TCV efficacy against infection of 15% to 85% in the primary model, and 60% to 90% in the alternative model. If adults had been vaccinated in the trial at the same coverage level as children, we estimate that indirect protection would have increased from 19% to between 40%-57%. These findings suggest that TCV confers protection against transmission from acute S. Typhi infections, and that the low indirect protection observed in the TyVAC-Bangladesh trial can be explained by transmission from unvaccinated adults.

## Introduction

Typhoid fever, a disease caused by *Salmonella enterica* serovar Typhi (S. Typhi), results in around 10 million cases and over 100 thousand deaths in low- and middle-income countries (LMICs) each year.(1,2) S. Typhi is transmitted through the fecal-oral route (usually contaminated food and water) and is almost exclusively found in settings with limited access to clean water, sanitation, and hygiene (WaSH) infrastructure. The burden of typhoid fever is concentrated in Africa and South Asia and is at risk of growing substantially in the future as a result of climate change, rapid urbanization, and the spread of antimicrobial resistance.

The current generation of typhoid conjugate vaccines (TCVs), in which the Vi-polysaccharide antigen of S. Typhi is conjugated to a carrier protein to boost immunogenicity, is associated with greater protection than previous typhoid vaccines, with a single dose of Typbar-TCV (Bharat Biotech) estimated to provide 83% protection against typhoid fever over two years of follow-up.(3) Since TCV can be administered to infants as young as 6 months of age, it can be incorporated into routine childhood immunization programs and has the potential to address the high burden of disease among children in endemic settings. TCV was recommended by the World Health Organization (WHO) in 2018, and has been introduced into routine immunization programs in Bangladesh, Burkina Faso, Kenya, Liberia, Malawi, Nepal, Pakistan, Samoa, and Zimbabwe.(4–8)

While the protective efficacy of TCV against clinical typhoid fever has been well-established through multiple field trials, it is unclear if the vaccine prevents *S*. Typhi infection and can induce population-level “herd” protection, or if vaccination simply blocks progression to symptomatic disease. It is important to clarify this, as many of the mathematical models that are used to inform policy decisions assume a transmission-blocking mechanism.(9) Indirect protection was observed in a cluster-randomized trial of ViPS vaccine in Kolkata, India, where both adults and children were vaccinated (indirect protection: 44%, 95% CI: 2%-69%), but not in Karachi, Pakistan, where vaccination was limited to children (indirect protection: -10%, 95% CI: - 116%-44%).(10,11) In the TyVAC-Bangladesh cluster-randomized trial, in which children 9 months to 15 years in Dhaka were vaccinated with Typbar-TCV, indirect protection of unvaccinated individuals was weak and not statistically significant (indirect protection: 19%, 95% CI: -12%-41%). However, indirect protection was only a secondary endpoint in the trial, and consequently the study was not explicitly powered to measure it.(12)

The reasons underlying the weak indirect protection from TCV in the TyVAC-Bangladesh trial have not been fully explored. Conjugate vaccines against respiratory-transmitted bacterial pathogens, including *H. influenzae* type b (Hib), pneumococcus, and meningococcus, have all demonstrated a robust impact on transmission(13–15), although this pattern may not be generalizable to enteric pathogens. In an immunogenicity trial in India, Typbar-TCV recipients were 63% less likely than ViPS recipients to have serological evidence of *S*. Typhi infection in the two years following vaccination.(16) TCV recipients were also less likely than unvaccinated recipients to shed S. Typhi in stool samples in controlled human challenge studies(17). It is possible that indirect protection in the TyVAC-Bangladesh trial was diluted due to contamination from outside, i.e. transmission between individuals in vaccinated versus unvaccinated clusters. However, analyses of the inner 75%, 50% and 25% subpopulations of the clusters did not reveal significant changes in indirect protection.(18,19) Furthermore, only 64% of children from 9 months to 15 years of age were vaccinated in this trial, and older age groups were not eligible for vaccination, although recent analyses show high rates of seroincidence among adults in Dhaka.(20) Vaccinating a larger share of the population may be needed to achieve measurable herd protection. Lastly, previous modeling studies have suggested that the level of indirect protection may be limited if chronic carriers play an important role in driving transmission.(21)

A better understanding of the low indirect protection observed in the TyVAC-Bangladesh trial and its causes is needed to determine whether TCVs can have a meaningful impact on transmission when implemented at the population level, and if so, how these transmission effects can be maximized to control typhoid fever. In this study, we used two transmission dynamic models to simulate the TyVAC-Bangladesh trial, identify profiles of vaccine protection against infection and chronic carrier transmissibility which are consistent with trial observations, and evaluate the impact of strategies for increasing the population-level impact of vaccination.

## Methods

### Study Description

The TyVAC-Bangladesh study was a cluster-randomized trial of Typbar-TCV conducted in Dhaka, Bangladesh between April 2018 and October 2019. The study area was divided into 150 contiguous clusters, each of which was randomly allocated to receive either TCV or a control vaccine (Japanese encephalitis vaccine [JEV]) in a 1:1 ratio. Eligible and consenting participants between 9 months and <16 years of age received a single vaccine dose and were followed for an average of 17.1 months (n = 61,657 total vaccinated participants). The average vaccine coverage among age-eligible cluster residents was 64% during the study period. The primary endpoints of the trial were total protection against typhoid fever in TCV recipients relative to JEV recipients (29 vs 192 blood-culture-confirmed cases; total protection: 85%, 95% CI: 78%-90%) and overall protection against typhoid fever among all treatment and control cluster residents (144 vs 331 blood-culture-confirmed cases; overall protection: 57%, 95% CI: 45%-67%). Indirect protection against typhoid fever among non-vaccinated treatment and control cluster residents was a secondary trial endpoint (115 vs 139 blood-culture-confirmed cases; indirect protection: 19%, 95% CI: -12%-41%).

Prior to the TyVAC-Bangladesh trial, the Bangladesh arm of the Strategic Typhoid Alliance Across Asia and Africa (STRATAA) study was conducted in the same Mirpur area of Dhaka to characterize the local burden and epidemiology of enteric fever at this site.(22) As part of the STRATAA study, data from passive blood-culture surveillance, household census activities, and a healthcare utilization survey were used to calculate the age-specific adjusted incidence of symptomatic typhoid at the Mirpur study site. The STRATAA study also included a serologic survey, in which paired blood samples were collected and used to estimate the age-specific incidence of typhoidal *Salmonella* infections based on several antigen targets.(20) For this study, we focused on seroincidence estimates based on the HlyE antigen, due to its strong and specific association with acute enteric fever cases at the individual and population levels.(20,23) However, HlyE is expressed by both *S*. Typhi and *S*. Paratyphi; to isolate serologic responses to the former and exclude the latter, we multiplied seroincidence estimates by the age-specific fraction of *S*. Typhi among blood-culture-positive enteric fever cases in the STRATAA-Bangladesh study.

### Model description and calibration

Our primary analysis is based on a transmission dynamic model of typhoid infection and disease(21), which has previously been used to evaluate the impact and cost-effectiveness of typhoid vaccination and investigate the drivers of typhoid fever outbreaks.(9,24–27) In this model, individuals are born into a susceptible and previously unexposed class *S*_1_, and are infected and moved into the *I*_1_ class at rate λ (the force of infection). After an average of 4 weeks, an age-specific fraction of primary infections become long-term chronic carriers (C), while the remainder recover into the *R*_1_ class. Recovered individuals remain immune to reinfection for an average of 1 year before waning into the *S*_2_ class. Previously infected susceptible individuals can be reinfected and move to *I*_2_, but we assume reinfections are subclinical and do not lead to chronic carriage. We also explored an alternative model in which this process is repeated to independently track second, third, and subsequent infections (*I*_2_, *I*_3_, and *I*_4_ respectively, Figure S1). We assume that previously exposed individuals in *S*_2-4_ are fully susceptible to reinfection (*I*_*2-4*_) but do not develop chronic carriage, and that the symptomatic fraction *f*_*i*_ (estimated during model fitting) may decrease with each subsequent infection *i*. For both models, we assume that the mean duration of infection (in the *I* classes) and immunity (in the *R* classes) is independent of infection history. Finally, we assumed that acute primary infections (*I*_*1*_) and reinfections (*I*_*2-4*_) transmit to susceptible individuals at the same rate (β), while the transmissibility of chronic carriers is a fraction *r*_*C*_ of acute infections.

Within the model, we stratified the population into eight age groups: 0 to <9 months, 9 months to <5 years, 5 to <10 years, 10 to <15 years, 15 to <16 years, 16 to <30 years, 30 to <50 years, and 50+ years old. These age groups were selected to align with the aggregated age-specific (sero)incidence in the STRATAA study and the ages that were eligible for vaccination in the TyVAC-Bangladesh trial (9 months to <16 years).(18) For susceptible populations aged 0 to <5 and 5 to <10 years old, we estimated a relative risk of infection (*b*_*1*_ and *b*_*2*_, respectively) compared to susceptible individuals in older age groups. Models were run for an initial 100-year burn-in period, during which population demographics and transmission dynamics converged to a steady state, followed by a 2-year observation period during which incidence was calculated.

In our primary model, we simultaneously estimate the maximum likelihood parameter values (β, *b*_*1*_, *b*_*2*_, *f*_*1*_) by fitting the modeled symptomatic incidence to the adjusted clinical typhoid incidence from the STRATAA-Bangladesh study. For the alternative model, we estimate the transmission parameters (β, *b*_*1*_, and *b*_*2*_) by fitting the incidence of infection to the adjusted STRATAA HlyE seroincidence (described above), and then estimate the symptomatic fraction parameters (*f*_*1*_, *f*_*2*_, *f*_*3*_, and *f*_*4*_) by fitting the modeled symptomatic incidence to the adjusted STRATAA typhoid incidence. These estimates are constrained such that the symptomatic fraction *f* is either reduced or stays constant with each additional infection (*f*_*1*_ ≥ *f*_*2*_ ≥ *f*_*3*_ ≥ *f*_*4*_). The seroincidence data suggests that the incidence of infection is highest within the first few years of life even though typhoid fever incidence peaks at a later age. To account for this in the alternative model, we assume that the symptomatic fraction is lower amongst those aged 0 to <5 years relative to older populations. This relative risk of symptoms is estimated alongside the other symptomatic fraction parameters during the model-fitting process. We fit the primary and alternative models independently for values of *r*_*C*_ between 0% and 100% (relative to the transmissibility of acute infections) in 5% increments. The values of all fixed, varied, and estimated model parameters are listed in Table 1.

**Table 1:** Parameter values for the transmission dynamic model.

| Fixed Parameters: Both Models |  |  |
| --- | --- | --- |
| Parameter | Parameter | Parameter |
| Births per 1000 person-years | 18.23 per 1000 person-years | STRATAA Census Data |
| Net mortality + migration rate per 1000 person-years | Age [0-5) years: -3.72<br>Age [5-10) years: 1.83<br>Age [10-15) years: -2.01<br>Age [15-30) years: -1.95<br>Age [30-50) years: 2.83<br>Age ≥ 50 years: 8.24 | Fitted to STRATAA Census Data (see Figure S2) |
| Mean duration of acute infection | 4 weeks | Hornick *et al.*(28) |
| Mean duration of post-infection immunity | 1 year | Assumption |
| Fraction of primary infections which become chronic carriers | Age [0-15y): 0.3%<br>Age [15-30y): 1.5%<br>Age [30-50y): 6.6%<br>Age ≥ 50y: 9% | Ames *et al.*(29) |
| Primary Model: Fitted and Fixed Parameters |  |  |
| Parameter | Value | Source |
| Transmission rate (β) | 0.13 to 0.27<sup>#</sup> | Fitted to adjusted STRATAA-Bangladesh typhoid fever incidence data, conditional on *rC* |
| Relative risk of infection (vs age ≥ 10 years) | [0-5) year olds: 0.70 to 0.71<sup>#</sup><br>[5-10) year olds: 1.34 to 1.35<sup>#</sup> |  |
| Symptomatic fraction: primary infection* | 0.59 to 0.61<sup>#</sup> |  |
| Symptomatic fraction: reinfections | 0 | Assumption |
| Alternative Model: Fitted Parameters |  |  |
| Parameter | Value | Source |
| Transmission rate (β) | 0.26 to 0.30<sup>#</sup> | Fitted to adjusted STRATAA-Bangladesh seroincidence data, conditional on *rC* |
| Relative risk of infection (vs age ≥ 10 years) | [0-5) year olds: *b*<sub>1</sub> = 5.24<br>[5-10) year olds: *b*<sub>2</sub> = 2.62 |  |
| Symptomatic fraction: primary infection, age ≥ 5 years old* | 0.39 | Fitted to adjusted STRATAA-Bangladesh typhoid fever incidence data, conditional on *rC*, β, *b*<sub>1</sub>, and *b*<sub>2</sub> |
| Relative symptomatic fraction* | 2<sup>nd</sup> vs 1<sup>st</sup> infection: 1<br>3<sup>rd</sup> vs 2<sup>nd</sup> infection: 1<br>4<sup>th</sup> and later vs 3<sup>rd</sup> infection: 0.04<br>Age [0-5) years vs older: 0.14 |  |
| Varied Parameters |  |  |
| Parameter | Value |  |
| Infectiousness of chronic carriers relative to acute infections (*rC*) | Varied from 0% to 100% in increments of 5% |  |
| Vaccine protection against infection (*VE*<sub>infection</sub>) | Varied from 0% to 100% in increments of 5% |  |

\* parameter estimate constrained between 0 and 1 during fitting
<sup>#</sup> range of parameter estimates across $rC = 0\%$ to $100\%$

### Simulating vaccination in the TyVAC-Bangladesh trial

To incorporate vaccination in the model, we created a vaccinated counterpart for each compartment (*S*_*1*_ and *Sv*_*1*_, *I*_*1*_ and *Iv*_*1*_, etc). At the end of the burn-in period, a proportion of individuals in the eligible age groups are moved from each compartment to its vaccinated counterpart. Unless otherwise stated, we assumed a target vaccine coverage of 64% among those aged 9 months to <16 years, based on the average level over the duration of the trial.(18) To keep vaccine coverage constant over time in the eligible age groups, we vaccinated the same proportion of infants at 9 months of age. A similar effect was achieved in the TyVAC-Bangladesh trial by performing catch-up vaccine campaigns every 6 months. In the vaccinated compartments, the force of infection is reduced by a fraction *VE*_*infection*_, while the symptomatic fraction is reduced by a fraction *VE*_*progression*_, resulting in a “leaky” mechanism of vaccine protection.(30) Given the limited duration of the TyVAC-Bangladesh trial (17.1 months of follow-up on average), we assume no waning of vaccine protection over time in our primary analysis. As a sensitivity analysis, we considered a scenario in which the duration of vaccine protection follows an exponential distribution with an average duration of 5 years.

Using the fitted models, we simulated the TyVAC-Bangladesh trial for 441 unique combinations of *r*_*C*_ and *VE*_*infection*_, with values of each parameter ranging from 0% to 100% in increments of 5%. For each of these parameter combinations, we ran two models, one corresponding to the TCV-receiving treatment clusters and one corresponding to the control clusters. To account for the impact of contamination (transmission between those residing TCV and control clusters), we reduced vaccine coverage in the treatment cluster model by 10% from the target coverage level, and increased vaccine coverage in the control cluster model from 0% to 10%.

### Estimating population-level vaccine effects

To calculate the total protective effect of vaccination (total VE) from the model output, we compared the expected incidence of typhoid fever between eligible TCV and JEV recipients in the treatment and control clusters, respectively. We calculated the overall protective effect of vaccination (overall VE) by comparing the expected incidence of typhoid fever between all individuals in the treatment and control clusters. Finally, to calculate the indirect protective effect of vaccination (indirect VE), we compared the expected incidence of typhoid fever between the unvaccinated populations of the treatment and control clusters. In these calculations, we standardized modeled incidence by the target vaccine coverage and the age distribution of vaccinated, unvaccinated, and overall population. The full equations describing these calculations can be found in the supplementary methods.

We classified pairs of *r*_*C*_ and *VE*_*infection*_ values as inconsistent with the TyVAC-Bangladesh trial results if the projected total, overall, or indirect effects of vaccination did not fall within the 95% confidence intervals of the trial estimates (total VE: 78%-90%, overall VE: 45%-67%, and indirect VE: -12%-41%) for any value of *VE*_*progression*_ between 0% and 100%.

### Simulating counterfactual vaccine trials

To evaluate the sensitivity of the trial results to vaccine coverage and age eligibility, we modeled vaccine effects under three scenarios:

- Counterfactual #1: increasing vaccine coverage to 90% of eligible children aged ≥ 9 months to <16 years old without extending vaccination to older age groups
- Counterfactual #2: extending vaccination to all children and adults aged ≥ 9 months old at 64% coverage
- Counterfactual #3: extending vaccination and increasing coverage to 90% of all children and adults aged ≥ 9 months old

We evaluated each scenario for three parameter sets which reproduce the 19% indirect protection estimate from the trial, corresponding to low, medium, and high values of *VE*_*infection*_ and *r*_*C*_, for both the primary and alternative models. For each parameter set, we calibrated *VE*_*progression*_ to reproduce the 85% total protection observed in the trial (see Supplementary Methods).

To compare the relative efficiency of increasing vaccine coverage among children and extending vaccination to adults, we calculated the ratio of the incremental number of averted cases and administered doses for each of the coverage scenarios described above, relative to a corresponding strategy in which vaccination is more limited. Specifically, we evaluated the efficiency of increasing vaccine coverage from 0% to 64% and from 64% to 90% of children aged ≥ 9 months to <16 years old, and of expanding vaccination from children only to both children and adults at the same coverage level (64% and 90%).

All modeling and analysis was performed in R (v4.1.1). Code and data are available from http://github.com/pitzerlab/Typhoid-Indirect-Protection.

## Results

Both the primary and alternative models captured the magnitude and age pattern of symptomatic typhoid fever incidence observed in Dhaka in the STRATAA study, with incidence peaking in the 5-to <10-year-old age group and then declining through adulthood, although the primary model reproduced these trends slightly more accurately than the alternative model (Figure 1). In the primary model, which was fit exclusively to the clinical surveillance data, the initial increase in typhoid incidence with age during childhood reflects a higher risk of infection in children aged 5 to <10 years (Figure 1A-B). In the alternative model, where the incidence of infection was fit to the seroincidence data, the risk of infection decreases with age in early childhood, even as typhoid fever incidence increases (Figure 1C-D). In both models, the risk of infection plateaus through adolescence and adulthood, but typhoid fever incidence continues to decline with age as acquired immunity from prior infections reduces the risk of progression to symptomatic disease. The age-specific incidence of infection was 4-to 15-fold higher in the alternative model than the primary model (27-73 vs 5-9 per 100 person-years).

**Figure 1:**
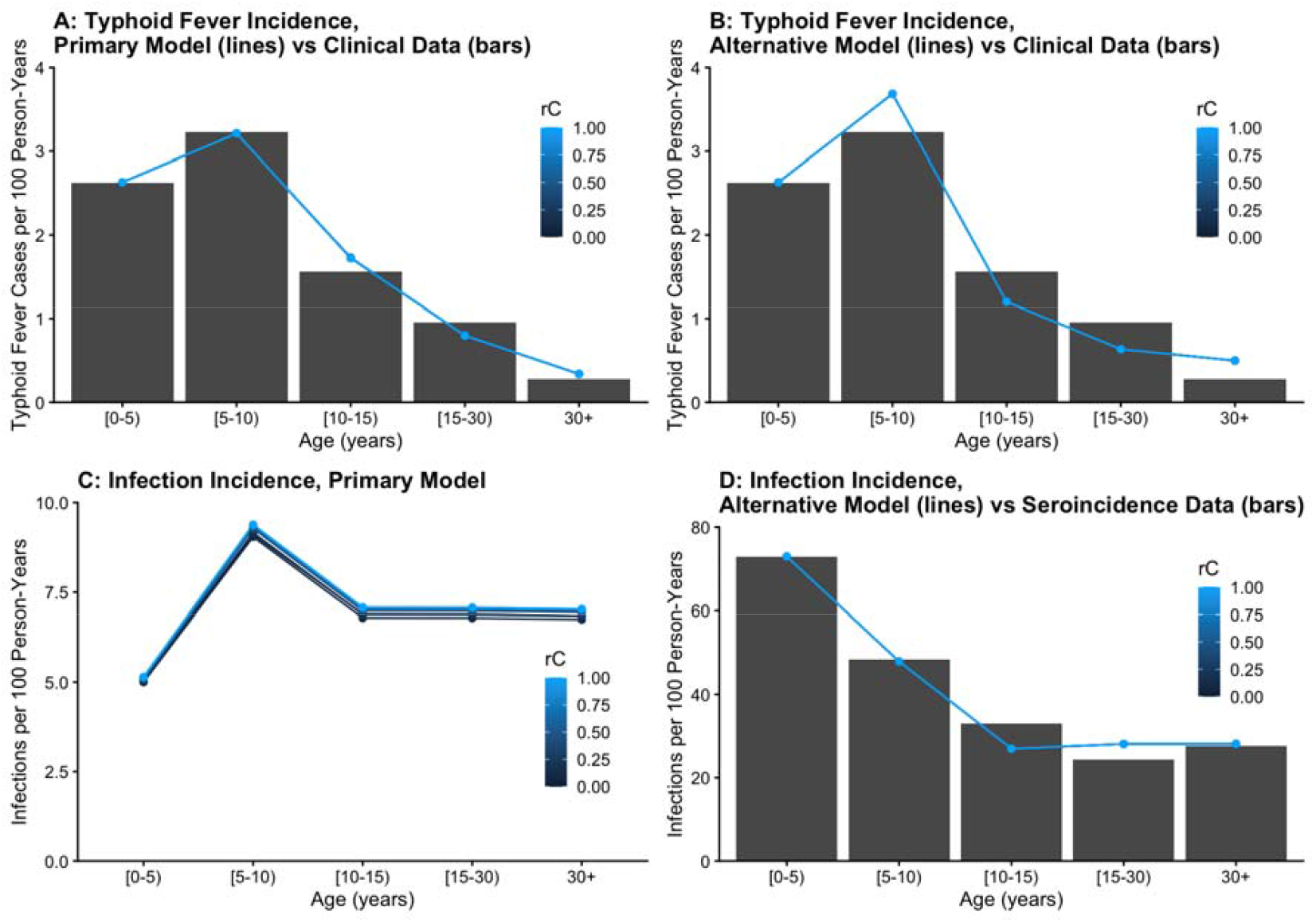
Estimated incidence of clinical typhoid fever incidence and seroincidence of *Salmonella* Typhi infection for the primary and alternative models. (A) Fitted typhoid fever incidence in the primary model (lines) against adjusted typhoid incidence in the STRATAA study (bars). (B) Fitted typhoid fever incidence in the alternative model (lines) against adjusted typhoid incidence in the STRATAA study (bars). (C) Estimated incidence of S. Typhi infection in the primary model. (D) Fitted incidence of S. Typhi infection in the alternative model and adjusted HlyE seroincidence in the STRATAA study.

In treatment clusters, the overall modeled incidence of typhoid fever ranged from 160 to 1,100 per 100,000 person-years in the primary model, and from 290 to 1,100 per 100,000 person-years in the alternative model, depending on the value of *r*_*C*_, *VE*_*infection*_, and *VE*_*progression*_. In control clusters, the overall modeled incidence of typhoid fever ranged from 800 to 1,100 per 100,000 person-years in the primary model, and from 1,000 to 1,100 per 100,000 person-years in the alternative model. The projected indirect protection ranged from 0% to 67% for the primary model, and from 0% to 54% for the alternative model, depending on the value of *VE*_*infection*_ and *r*_*C*_ (Figures 2 and 3). Indirect protection increased monotonically with *VE*_*infection*_ and decreased monotonically with *r*_*C*_ when holding the other parameter constant. For both the primary and alternative models, we identified a set of *VE*_*infection*_ and *r*_*C*_ values which were consistent with the TyVAC-Bangladesh trial results (Figure 3, Figures S3-S6), in that the modeled total, overall, and indirect effects of vaccination all overlapped with the 95% confidence interval of the trial estimates for some range of non-negative *VE*_*progression*_ values. The values of *VE*_*infection*_ that were consistent with the trial results ranged from 15% to 85% for the primary model, and from 60% to 90% for the alternative model, although this range was narrower for specific values of *r*_*C*_. Of these qualifying parameters, the values of *VE*_*infection*_ that most closely reproduced the 19% indirect protection observed in the trial ranged from 15% to 85% in the primary model (for *r*_*C*_ = 0% and 35%, respectively), and from 60% to 75% in the alternative model (for *r*_*C*_ = 0% and 100%, respectively). In sensitivity analyses, modeled indirect protection was up to 16% higher if there is no contamination across cluster boundaries, and up to 15% lower if TCV-induced immunity wanes after an average of five years (see Supplementary Findings).

**Figure 2:**
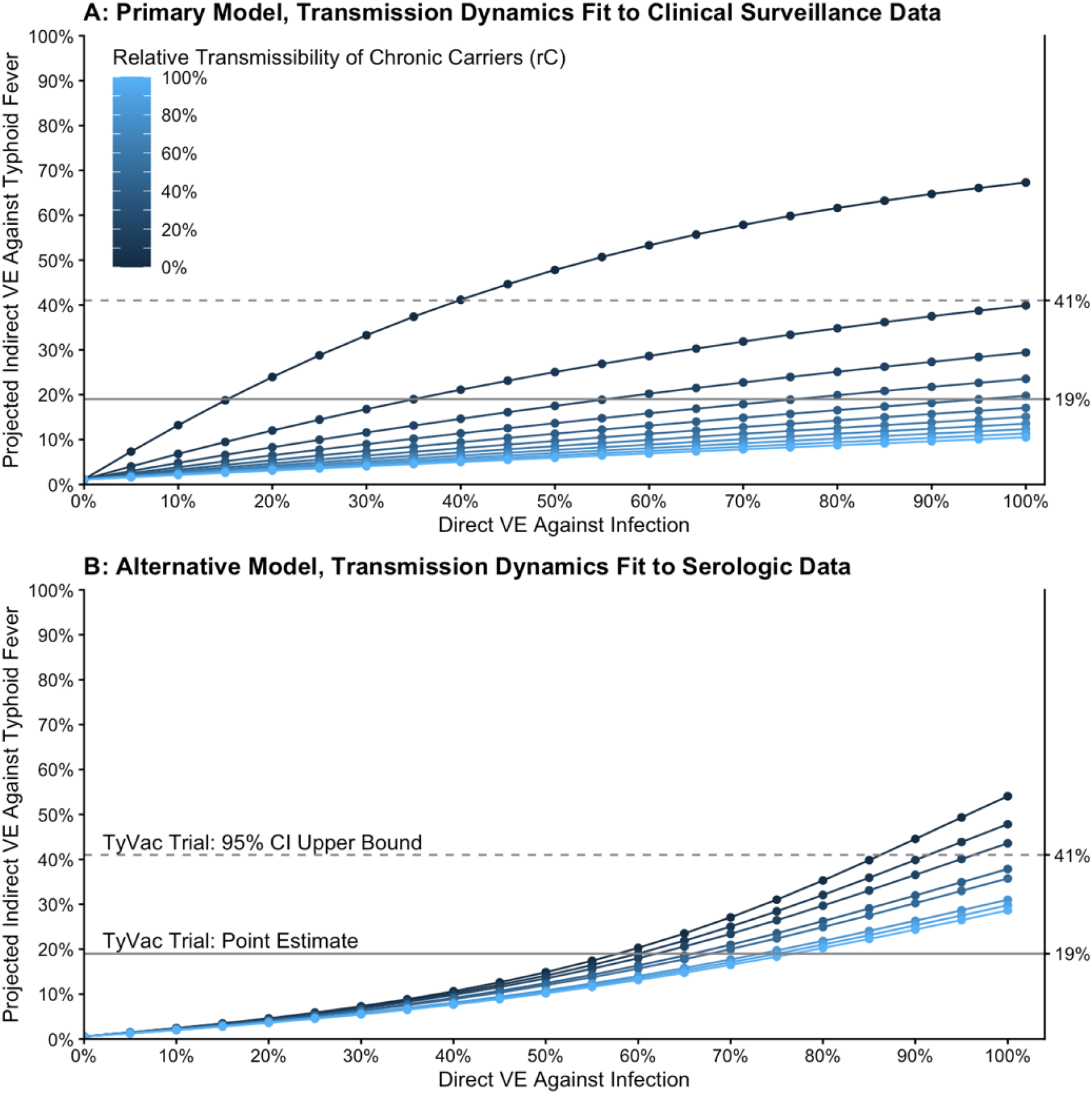
Model-predicted indirect protection for different levels of direct protection against infection and relative transmissibility of chronic carriers. Each line represents the model-predicted indirect protection (y-axis) under simulated cluster-randomized trial conditions mimicking the TyVAC-Bangladesh trial as a function of TCV efficacy against infection, with lighter colored lines corresponding to higher chronic carrier transmissibility. The top (A) and bottom (B) panels correspond to the primary and alternative models, respectively. The solid and dashed horizontal line represents the point estimate (19%) and 95% confidence interval upper bound (41%) of indirect protection in the TyVAC-Bangladesh trial.

**Figure 3:**
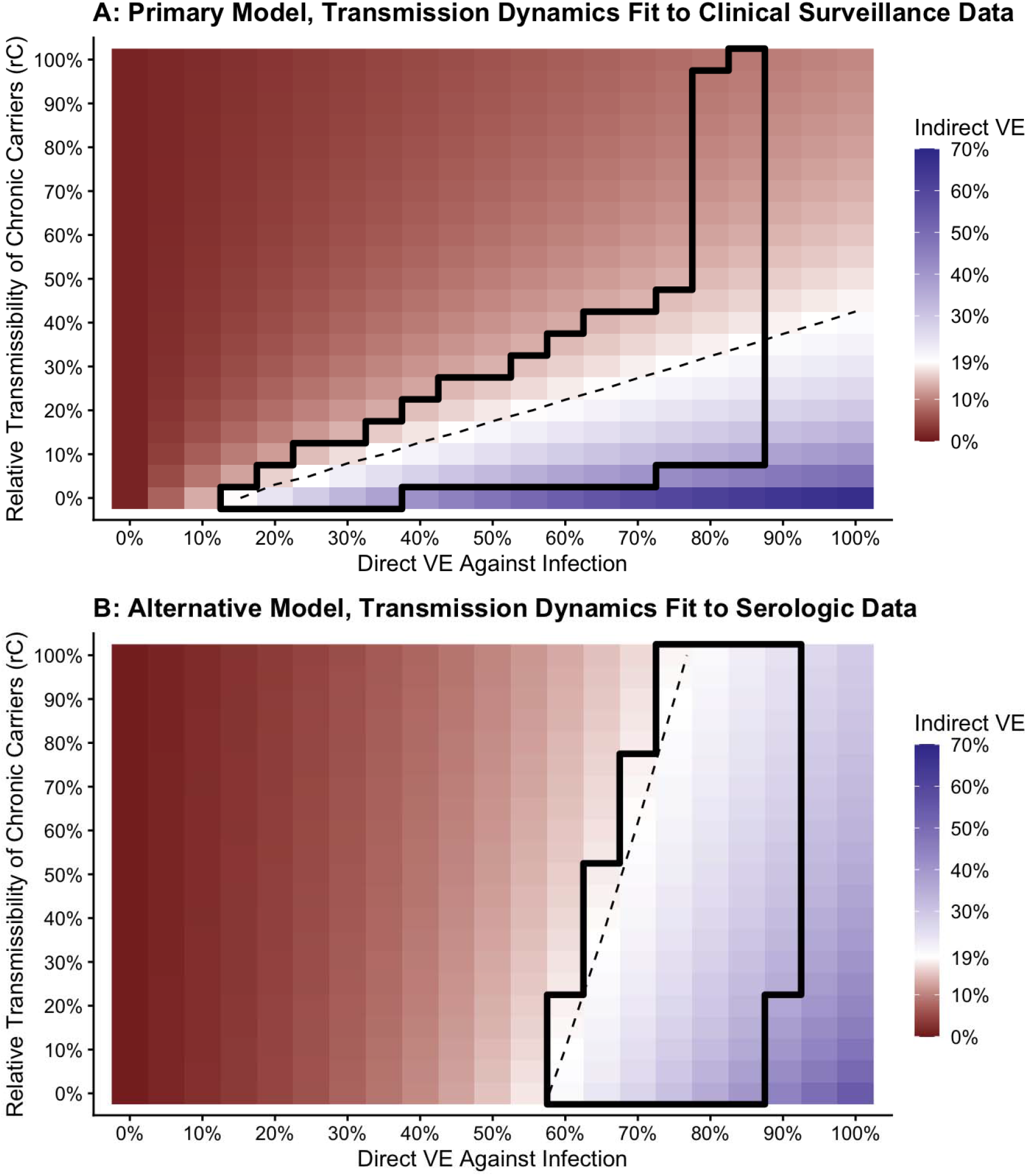
Heatmap of the model-predicted indirect protection for different levels of direct protection against infection and relative transmissibility of chronic carriers. Each pixel represents the projected indirect protection in the TyVAC-Bangladesh trial for a given level of TCV efficacy against *S*. Typhi infection (x-axis) and relative transmissibility of chronic carriers compared to acute cases (y-axis). The region bounded by the solid black line contains parameter combinations for which model outputs are consistent with the 95% confidence intervals of the total, overall, and indirect protection in the TyVAC-Bangladesh trial. The dashed line interpolates parameter sets which reproduce the 19% trial estimate of indirect protection. The top (A) and bottom (B) panels correspond to the primary and alternative models, respectively.

If the 85% total protection and 19% indirect protection estimate from the TyVAC-Bangladesh trial represent the “true” effects under the observed conditions, we estimate that increasing vaccine coverage to 90% of eligible children would have resulted in 28%-29% indirect protection and 62% overall protection in the primary model, and 33%-38% indirect protection and 62%-64% overall protection in the alternative model, depending on the values of *VE*_*infection*_ and *r*_*C*_ (Figures 4-5). Expanding vaccination to adults at the same 64% coverage level achieved in the trial had an even greater impact, resulting in 40%-47% indirect projection and 70%-73% overall protection in the primary model, and 42%-57% indirect projection and 71%-79% in the alternative model. Combining these effects by increasing vaccine coverage to 90% in children and adults amplified their impact, with a projected 52%-61% indirect protection and 86%-89% overall protection in the primary model, and 59%-80% indirect protection and 89%-94% overall protection in the alternative model. However, on a per-dose basis, increasing vaccine coverage in children was more efficient at preventing cases than expanding vaccination to adults (44-57 vs 9-17 averted cases per 1,000 doses over two years of follow-up, Table 2). The impact of expanding vaccine eligibility and increasing coverage beyond that observed in the TyVAC-Bangladesh trial was greatest for lower values of *rC*.

**Table 2:** Efficiency of increasing vaccine coverage in children and extending vaccination to adults.

| Model | Parameter Set | Incremental Averted Cases per<br>Additional 1,000 Doses Administered |  |  |  |
| --- | --- | --- | --- | --- | --- |
|  |  | Increasing<br>Vaccine<br>Coverage in<br>Children from<br>0% to 64%<br>(No Adult<br>Vaccination) | Increasing<br>Vaccine<br>Coverage in<br>Children from<br>64% to 90%<br>(No Adult<br>Vaccination) | Extending<br>Vaccination<br>to Adults<br>Alongside<br>Children:<br>64%<br>Coverage | Extending<br>Vaccination<br>to Adults<br>Alongside<br>Children:<br>90%<br>Coverage |
| Primary<br>Model | Low $VE_{infection}$ (15%)<br>and $r_c$ (0%) | 57 | 45 | 14 | 9 |
| | Medium $VE_{infection}$ (46%)<br>and $r_c$ (15%) | 57 | 44 | 13 | 9 |
| | High $VE_{infection}$ (76%)<br>and $r_c$ (30%) | 57 | 44 | 13 | 9 |
| Alternative<br>Model | Low $VE_{infection}$ (60%)<br>and $r_c$ (10%) | 54 | 59 | 17 | 10 |
| | Medium $VE_{infection}$ (68%)<br>and $r_c$ (50%) | 54 | 54 | 15 | 10 |
| | High $VE_{infection}$ (77%)<br>and $r_c$ (100%) | 54 | 52 | 14 | 9 |

**Figure 4:**
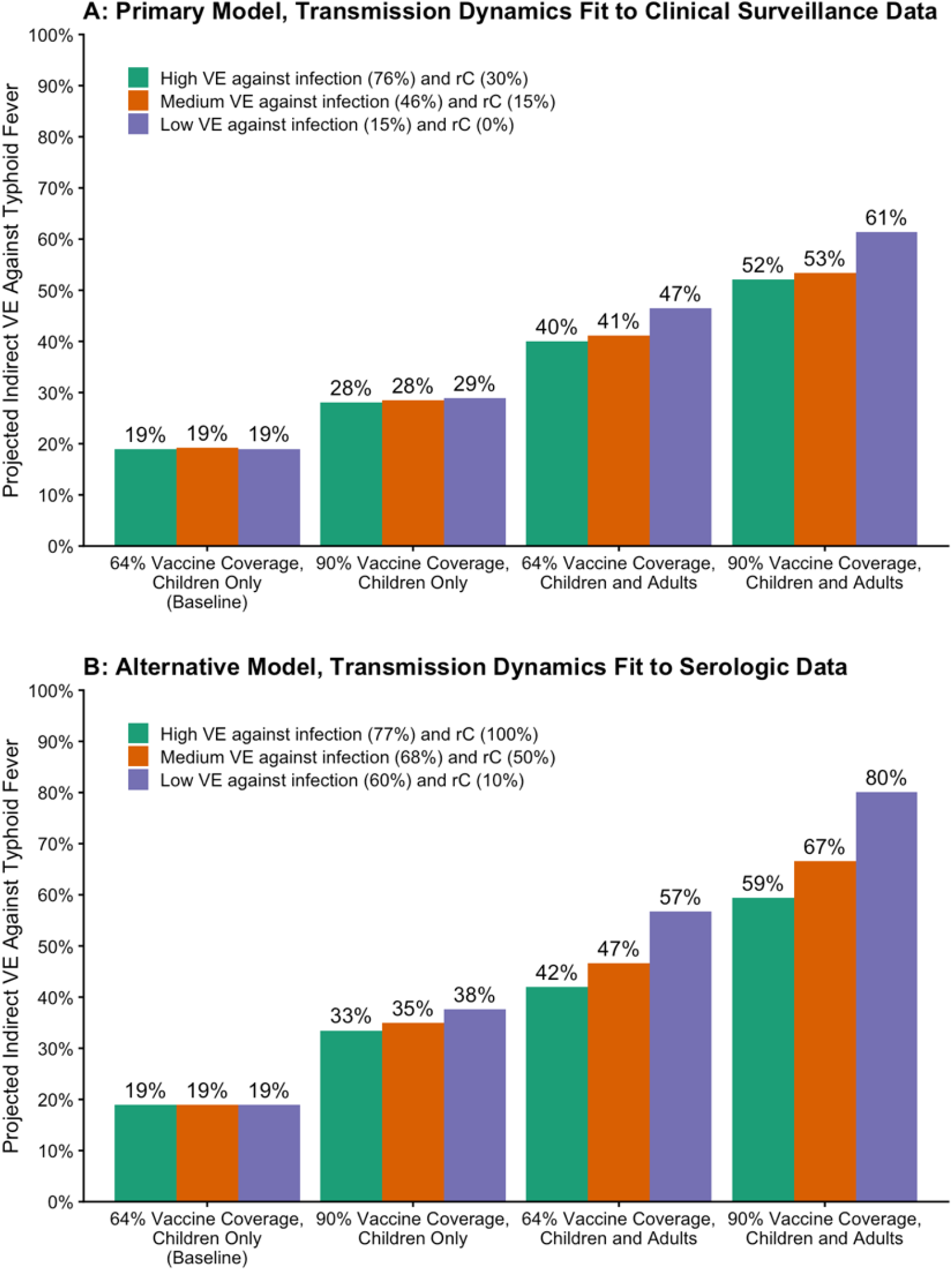
Projections of indirect protection under alternative trial conditions. Model-based projections of indirect protection under TyVAC-Bangladesh trial conditions (baseline) and in alternative scenarios in which vaccine eligibility is expanded to adults, vaccine coverage in the eligible population is increased from 64% to 90%, or both. Parameter sets are selected to reproduce the 19% indirect protection observed in the TyVAC-Bangladesh trial (Baseline).

**Figure 5:**
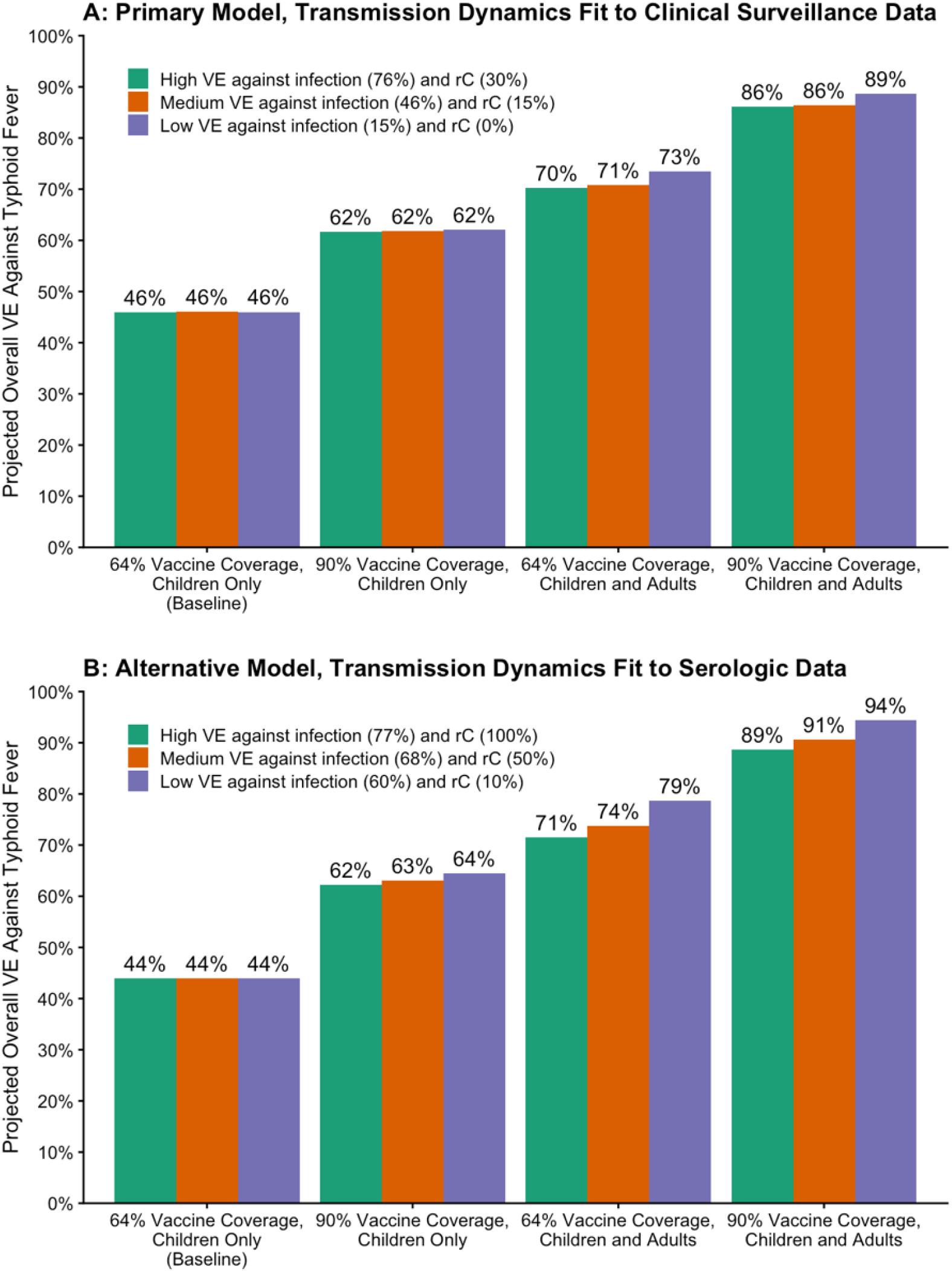
Projections of overall protection under alternative trial conditions. Model-based projections of overall protection under TyVAC-Bangladesh trial conditions (baseline) and in alternative scenarios in which vaccine eligibility is expanded to adults, vaccine coverage in the eligible population is increased from 64% to 90%, or both. Parameter sets are selected to reproduce the 19% indirect protection and 85% total protection observed in the TyVAC-Bangladesh trial (Baseline).

## Discussion

Typhoid conjugate vaccines confer strong protection against typhoid fever, as observed in multiple randomized control trials(3,18,31,32), human challenge studies(33), and observational studies.(34–36) However, the absence of significant indirect protection in the TyVAC-Bangladesh cluster-randomized trial raises the question of whether TCV is effective at preventing transmission and can induce meaningful herd effects when introduced more broadly. By simulating the trial with a mathematical model, we found that the low indirect protection observed in the TyVAC-Bangladesh trial does not necessarily imply low TCV efficacy against transmission. In both our primary and alternative models, vaccines which did not confer any protection against S. Typhi infection and transmission were inconsistent with the results of the TyVAC-Bangladesh trial, because overall protection would have been too low. On the other hand, scenarios in which TCV confers strong direct protection against infection were plausible in both models for a wide range of carrier transmission assumptions.

Our analysis suggests that the ineligibility of adults for vaccination was the primary explanation for low indirect protection in the TyVAC-Bangladesh trial, with moderate vaccine coverage among eligible children and the potential for transmission across cluster boundaries (i.e. contamination) being secondary drivers. It is important to emphasize that these factors have practical explanations, and do not represent failures of trial design or execution. Vaccination in the TyVAC-Bangladesh trial was restricted to children <16 years old, since these age groups account for most of the burden of disease in Bangladesh and other typhoid-endemic settings.(37) For a clinical trial of a relatively new vaccine in a setting with high levels of migration, achieving 64% coverage in eligible children required extensive planning, community engagement, and multiple rounds of immunization.

It is plausible that herd effects will be stronger than they were in the TyVAC-Bangladesh trial when TCVs are rolled out in the general population, at least temporarily. Immunization coverage could be higher for a proven vaccine in a non-trial setting, and recent nationwide TCV campaigns in Malawi and Nepal have achieved 77% and >95% coverage, respectively, based on administrative data.(5,38) Furthermore, the effect of migration and transmission from non-vaccinated areas would be minimized if TCV is introduced over a large geographic region, rather than in small neighborhood-level clusters. While not currently the WHO-recommended strategy, stronger herd effects could be achieved by vaccinating adults. Specifically, if adults had been vaccinated in the TyVAC-Bangladesh trial at the same coverage level as children, we estimate that indirect protection would have increased to between 40% and 64% depending on modeling assumptions. This effect is comparable to or greater than the 44% indirect protection in the Kolkata cluster-randomized trial of ViPS, in which children and adults were vaccinated and total protection was only 61%.(10) However, vaccination of adults in endemic areas would not necessarily be cost-effective or a public health priority, since the burden of clinical disease is concentrated in children: in our analysis, vaccinating children was over three times more efficient at preventing typhoid fever cases than vaccinating adults, on a per-dose basis. In addition, the waning of TCV-induced immunity could erode the strength of indirect protection over time. In a test-negative study performed in the years following the initial TyVAC-Bangladesh trial, TCV protection against typhoid fever declined from 93% (95% CI: 86%-96%) to 39% (95% CI: -23%-70%) between the first and fifth year post-vaccination.(39) The durability of direct and indirect protection from TCV and the cost-effectiveness of including adults in TCV campaigns should be evaluated in future modeling analyses.

A key observation, which has been described in previous modeling analyses, is that indirect vaccine protection is expected to be lower when chronic carriers drive a greater share of population-level transmission.(21) While vaccination can prevent new chronic infections, chronic carriers who were infected before vaccination began may continue to shed bacteria for years or even decades. As a result, the size of the chronic carrier population is expected to decline very slowly in response to vaccination. By contrast, acute infections generally recover after a few weeks, so a reduction in incidence quickly translates into a smaller infectious population. Mathematical models of typhoid fever transmission should account for uncertainty in the importance of chronic carriers to endemic transmission. In our analysis, we accomplish this by varying the relative transmissibility of chronic carriers while holding their prevalence fixed at a low level predicted by the model (0.6% and 0.3% in the primary and alternative models, respectively, in the absence of vaccination, see Figure S7). These modeled estimates of chronic carrier prevalence are consistent with the finding of one stool-culture-positive S. Typhi carrier among 192 healthy serosurvey participants with high IgG titers against the Vi antigen (0.52%), a potential marker of chronic carriage.(40–42) Previous modeling analyses predicting the impact and cost-effectiveness of TCV strategies in Gavi-eligible countries sampled *r*_*C*_ from an uncertainty distribution with an expected value of 25%, which is consistent with the TyVAC-Bangladesh trial results.(24,43)

Our analysis also demonstrates that models of TCV impact are sensitive to the mechanism of direct protection. In transmission dynamic models, vaccine effects often derive entirely from immunity to infection, potentially resulting in overly optimistic levels of herd protection, while static cohort models generally do not account for indirect effects at all. Model comparison studies which include both approaches can be a useful way to evaluate uncertainty in the potential impact of vaccine policies.(9) Transmission dynamic models should be validated against (or calibrated to) observed data on vaccine impact prior to making predictions.

In our model, we assumed vaccine uptake was uniform across the eligible population and therefore independent of the baseline risk of typhoid fever. However, this may not have been the case in the TyVAC-Bangladesh trial, where typhoid incidence in the control clusters was higher among control vaccine recipients than vaccine-eligible non-recipients (616 vs 406 cases per 100,000 person-years in those aged 2 to <16 years).(18) Residents who agree to participate in a vaccine trial may also be more likely to seek medical care for typhoidal illness.(44) Regardless, vaccine efficacy estimates from the trial should be robust to these differences and comparable to our model-based vaccine effect estimates for several reasons. First, when estimating total and indirect effects, the TyVAC-Bangladesh trial design inherently controls for any association between vaccine acceptance and the baseline typhoid risk, because the analysis is restricted to those who either did or did not accept the vaccine. Second, in the statistical analysis of the TyVAC-Bangladesh trial data, estimates of vaccine efficacy were adjusted for several factors that could influence care-seeking and the baseline risk of typhoid fever, including age, sex, reported handwashing behavior, the source and treatment of household drinking water, and household toilet type.

Our model simplifies the process of *S*. Typhi transmission by assuming homogenous mixing, in which susceptible and infected individuals come into contact at random. In reality, transmission-relevant contacts may be shaped by household structure and patterns of food consumption outside of the home, although indirect transmission through contaminated water sources may have a more generalized exposure pattern. In addition, our results are specific to the Dhaka, Bangladesh study site and should not be over-interpreted or extrapolated to other settings.

Our findings suggest that TCVs confer protection against *S*. Typhi infection and transmission, not just symptomatic disease, and that the low indirect protection observed in the TyVAC-Bangladesh study can be explained by subclinical transmission from unvaccinated adults. Mathematical models of typhoid fever vaccination should explicitly account for indirect protection and the observed uncertainty in its magnitude. Bangladesh is planning to introduce TCV nationally later in 2025 by implementing routine vaccination at 9 months of age following a nationwide catch-up campaign in children between 9 months and 15 years old. Our analysis suggests that this strategy could at least temporarily induce moderate levels of indirect protection, depending on coverage and the duration of vaccine protection. Routine surveillance programs are needed to evaluate the population-level impact of TCVs and inform policy decisions in Bangladesh and other settings that introduce the vaccine. Even modest indirect effects would increase the overall impact of TCV introduction, making it an even more attractive tool for preventing typhoid fever.

## Data Availability

Code and data are available online at github.com/pitzerlab/Typhoid-IndirectProtection.

https://www.github.com/pitzerlab/Typhoid-IndirectProtection

## Supplementary Material

### Supplementary Methods: Calculating Vaccine Effects

Given the premise that JEV confers no protection against typhoid fever, we assumed that the modeled age-specific incidence was equivalent between unvaccinated residents and JEV recipients in the control cluster.

#### Total Protection

To calculate the total protective effect of vaccination (total VE), we compared the expected incidence of typhoid fever between eligible TCV and JEV recipients in the treatment and control clusters, respectively.

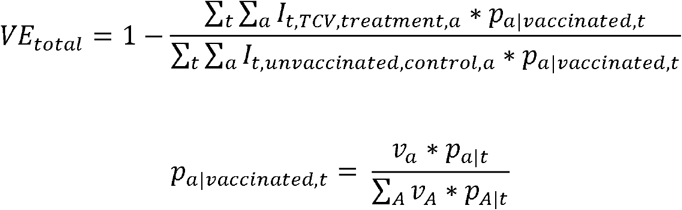

where

- *I*_*t,v,c,a*_ is the modeled per-capita incidence of typhoid fever in week *t* given vaccine status *v* (either *vaccinated* with TCV or *unvaccinated*), cluster *c* (*treatment* or *control*), and age group *a*.
- *p*_*a*|*vaccinated,t*_ is the proportion of the vaccinated population in age group a during week *t*
- *v*_*a*_ is the target vaccine coverage in age group *a*.
  ∘ When simulating the TyVAC-Bangladesh trial, we assumed *v*_*a*_ = 0.64 in children aged 9 months to <16 years old, and *v*_*a*_ = 0 for all other age groups, based on the observed coverage and eligibility criteria.
  ∘ In simulations of alternative trial scenarios, we explored the impact of increasing *v*_*a*_ to up to 90% of children and adults above 9 months of age.
- *p*_*a*|*t*_is the proportion of the overall model population in age group *a* during week *t*

#### Overall Protection

To calculate the overall protective effect of vaccination (overall VE), we compared the expected incidence of typhoid fever between all individuals in the treatment and control clusters:

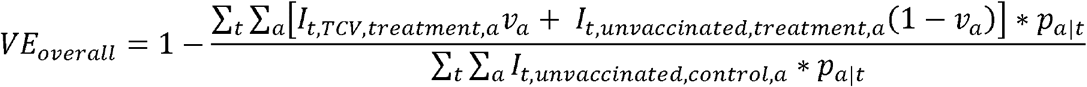

#### Indirect Protection

Finally, to calculate the indirect protective effect of vaccination (indirect VE), we compared the expected incidence of typhoid fever between the unvaccinated populations of the treatment and control clusters:

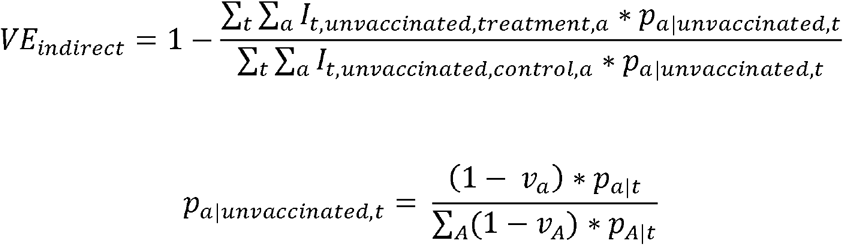

where *p*_*a*|*unvaccinated,t*_ is the proportion of the unvaccinated population in age group *a* during week *t*.

### Supplementary Methods: Calibrating Vaccine Protection Against Progression to Symptomatic Disease

Total protection *T* is a function of direct protection against infection and progression to symptomatic disease (*VE*_*infection*_ and *VE*_*progression*_, respectively), and indirect protection against infection:

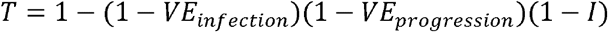

where *T* is total protection, *I* is indirect protection, and *VE*_*infection*_ and *VE*_*progression*_ are direct protection against infection and progression to symptomatic disease, respectively.

The combined effect of direct and indirect protection against infection can be obtained by estimating total protection in the model when *VE*_*progression*_ is set to 0:

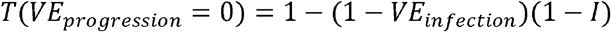

Once this “baseline” level of total protection is known, the total protective effect can be calculated analytically for any value v of *VE*_*progression*_ between 0 and 1:

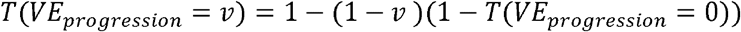

After substituting 0.85 (the TyVAC-Bangladesh trial estimate of total protection) for *T*(*VE*_*progression*_ = *v*) and the modeled total effect when *VE*_*progression*_ is set to 0 for *T*(*VE*_*progression*_ = 0), this equation can be rearranged to solve for the calibrated value of *VE*_*progression*_:

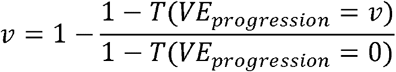

### Supplementary Findings: Alternative Contamination and TCV Waning Scenarios

In the primary analysis, we accounted for the effect of contamination (transmission across cluster boundaries), which is expected to reduce differences in incidence between treatment and control clusters and bias estimates of vaccine protection toward the null (0%), by reducing TCV coverage by 10% (from 64% to 54%) in the treatment clusters and increasing TCV coverage by 10% (from 0% to 10%) in the control clusters. As a sensitivity analysis, we simulated the TyVAC-Bangladesh trial while assuming no contamination across cluster boundaries (no adjustments to TCV coverage). Eliminating contamination increased the projected level of indirect protection by up to 13% and 16% for the primary and alternative models, respectively, and shifted the parameter sets which were most consistent with the trial estimates towards lower values of *VE*_*infection*_ and higher values of *r*_*C*_ (Figure S8).

Another assumption we made in the primary analysis is that TCV-induced immunity is long-lasting and did not meaningfully wane over the two-year follow-up period in the model. As a sensitivity analysis, we considered a scenario in which the duration of TCV protection only lasts for five years on average, and follows an exponential distribution. This level of waning reduced indirect protection by up to 4% and 15% in the primary and alternative models, respectively, and shifted the best-fitting parameter sets toward higher values of *VE*_*infection*_ and lower values of *r*_*C*_ (Figure S9).

**Supplementary Figure 1:**
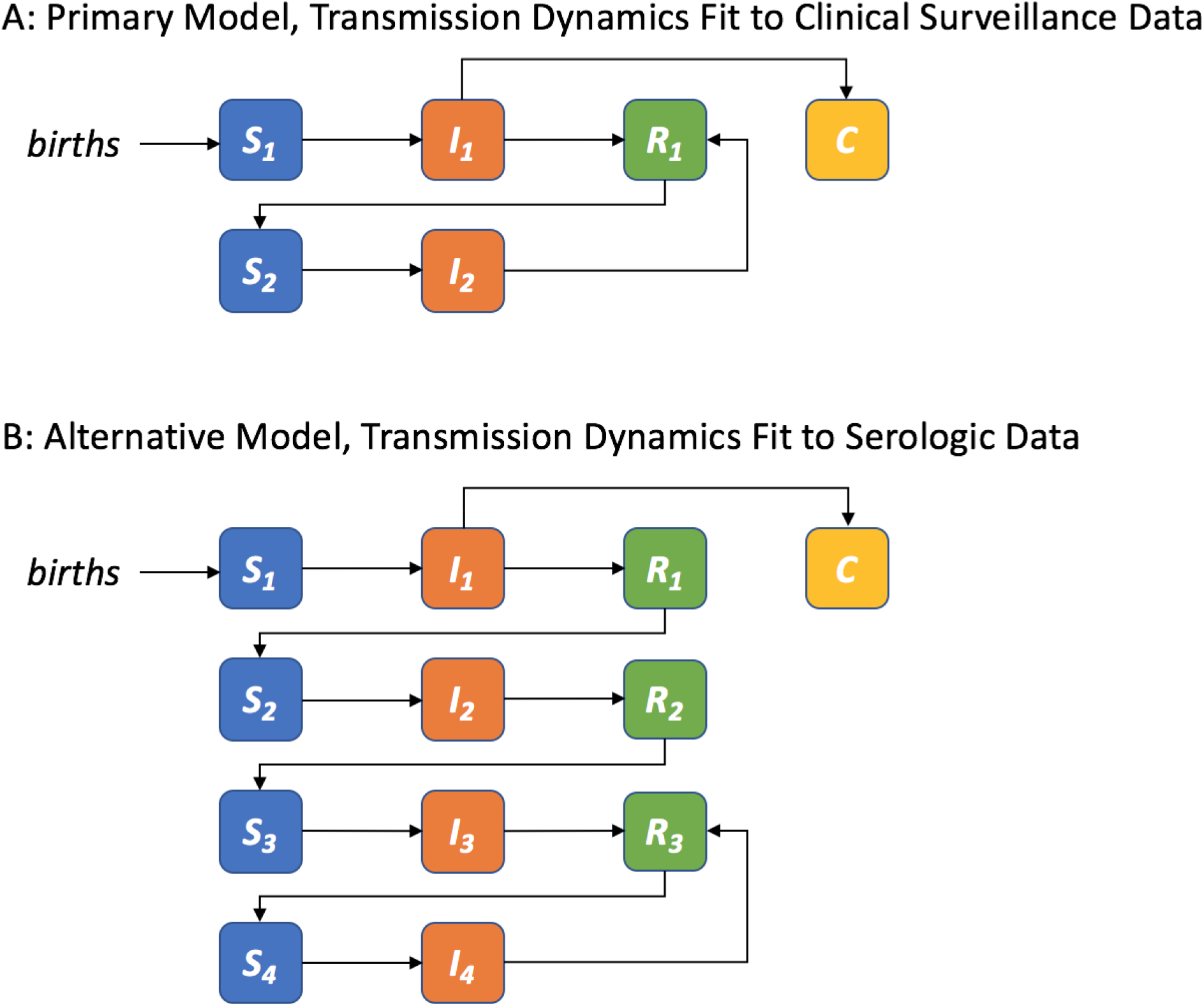
Diagram of the transmission model structure. Each box represents a distinct class that population members move between over time. The susceptible population remains in one of the blue *S* classes until they are infected with *S*. Typhi and move to an orange *I* class. In the primary model (A), an estimated fraction of primary infections become symptomatic typhoid fever cases, while all reinfections (*I*_*2*_) are assumed to be asymptomatic. In the alternative model (B), first, second, third, and subsequent infections (*I*_*1*_, *I*_*2*_, *I*_*3*_, and *I*_*4*_ respectively) are modeled separately, each with a corresponding symptomatic fraction. Infected individuals move to one of the green *R* classes after recovering, where they remain immune to reinfection until they wane back into a susceptible class. A fraction of primary infections (*I*_*1*_) do not recover, and become lifelong chronic carriers in the yellow *C* class. These classes are duplicated and run in parallel for each age group and vaccine status combination. Population members are moved to parallel set of classes when they are vaccinated or age into an older age group. Individuals are born into the susceptible *S*_*1*_ class of the youngest age group, and individuals that die are removed from the model (not pictured) at an age-specific mortality rate.

**Supplementary Figure 2:**
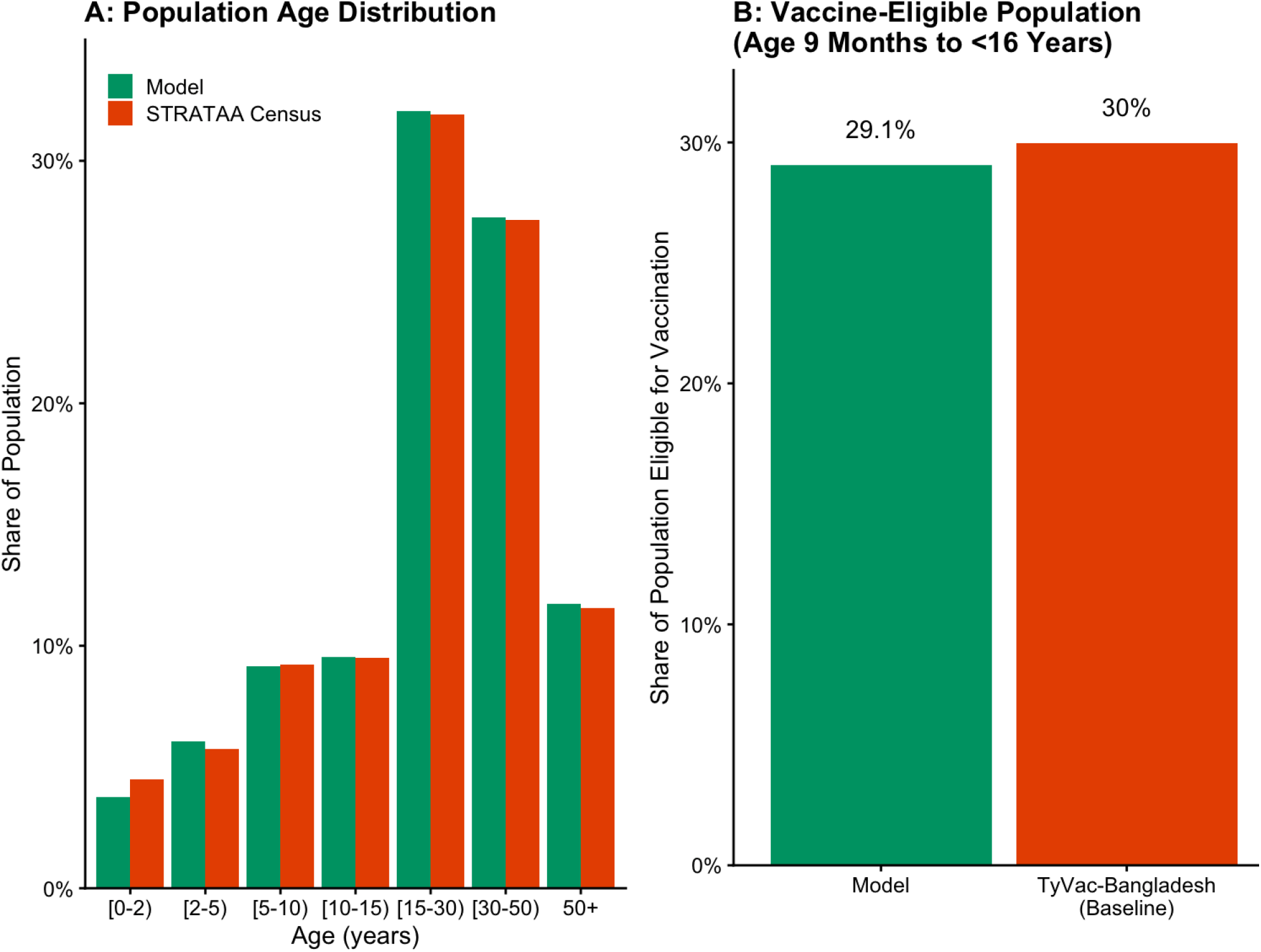
Modeled population age distribution. (A) The fraction of the population in each age group, in model simulations (green) and household census data from the STRATAA study (orange). (B) The fraction of the population which is eligible for vaccination (9 months to <16 years old) in the model and in STRATAA household census data.

**Supplementary Figure 3:**
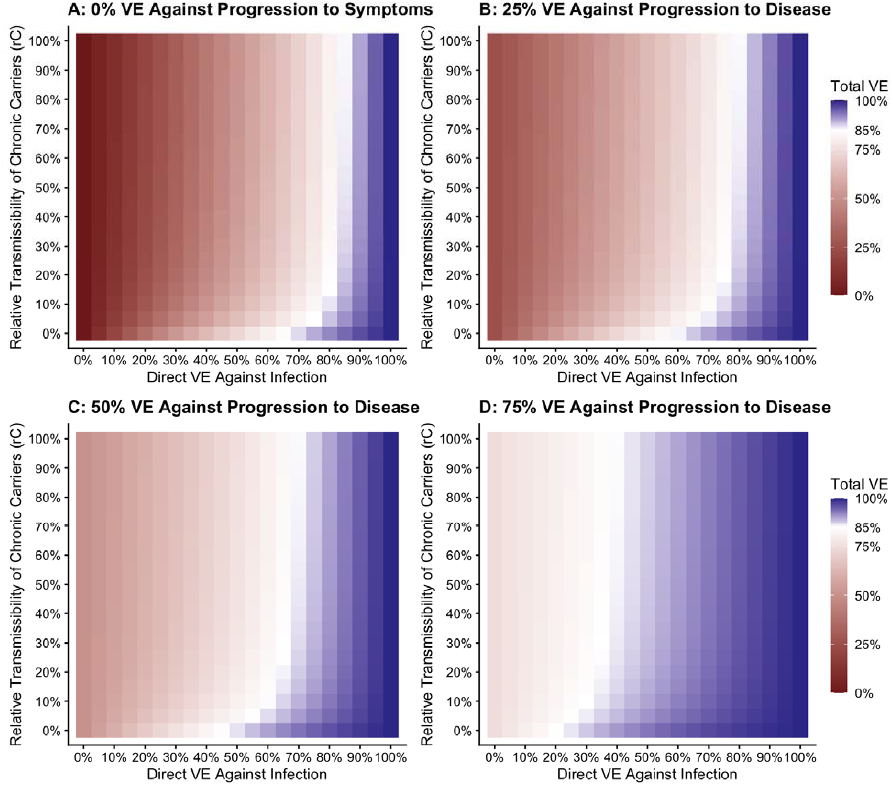
Projected total trial effect by direct vaccine efficacy against infection and the transmissibility of chronic carriers, primary model. This figure demonstrates the impact of *VE*_*progression*_ on total vaccine protection in the primary model. Each pixel represents the projected total protection in the TyVAC-Bangladesh trial for a given level of TCV efficacy against *S*. Typhi infection (x-axis) and relative transmissibility of chronic carriers compared to acute cases (y-axis). In each panel, TCV efficacy against progression from infection to symptomatic disease is assumed to be either 0% (A), 25% (B), 50% (C), or 75% (D).

**Supplementary Figure 4:**
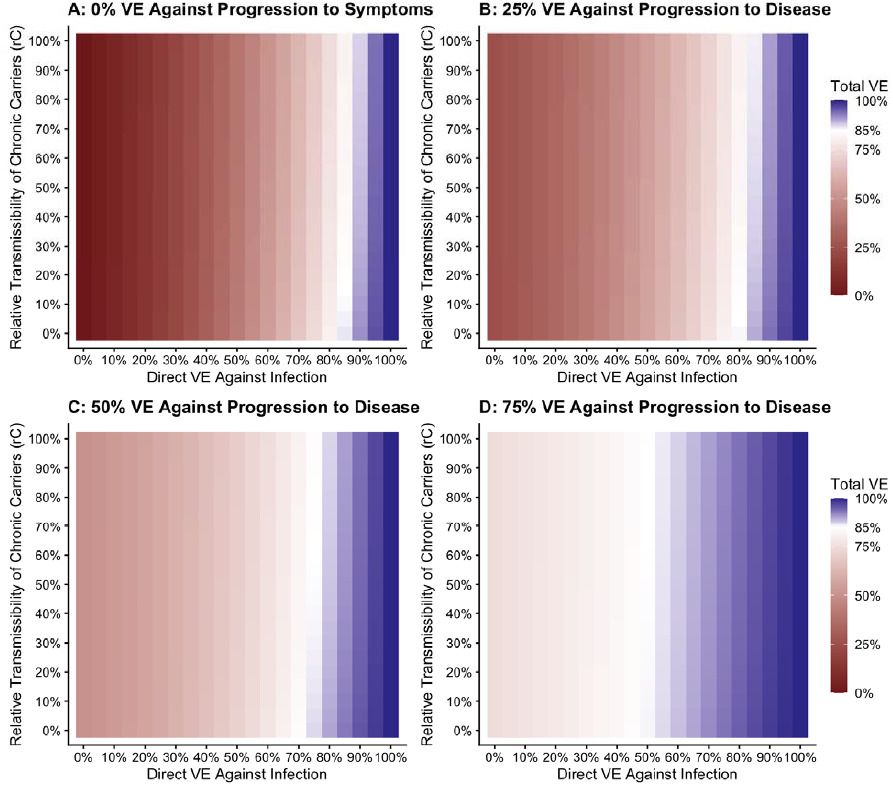
Projected total trial effect by direct vaccine efficacy against infection and the transmissibility of chronic carriers, alternative model. This figure demonstrates the impact of *VE*_*progression*_ on total vaccine protection in the alternative model. Each pixel represents the projected total protection in the TyVAC-Bangladesh trial for a given level of TCV efficacy against *S*. Typhi infection (x-axis) and relative transmissibility of chronic carriers compared to acute cases (y-axis). In each panel, TCV efficacy against progression from infection to symptomatic disease is assumed to be either 0% (A), 25% (B), 50% (C), or 75% (D).

**Supplementary Figure 5:**
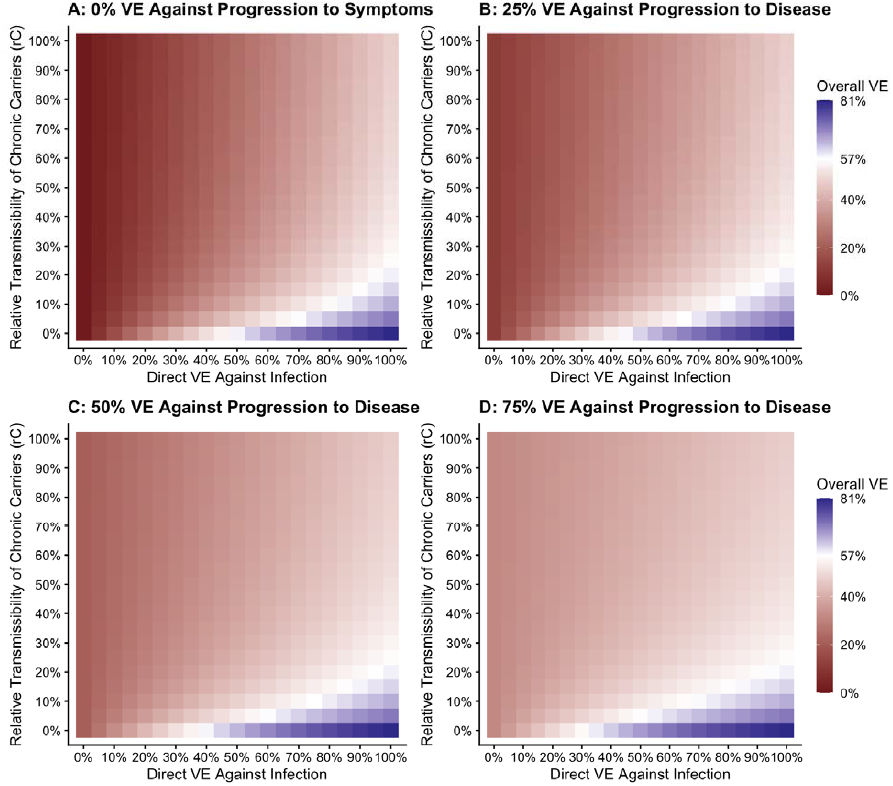
Projected overall trial effect by direct vaccine efficacy against infection and the transmissibility of chronic carriers, primary model. This figure demonstrates the impact of *VE*_*progression*_ on overall vaccine protection in the primary model. Each pixel represents the projected overall protection in the TyVAC-Bangladesh trial for a given level of TCV efficacy against *S*. Typhi infection (x-axis) and relative transmissibility of chronic carriers compared to acute cases (y-axis). In each panel, TCV efficacy against progression from infection to symptomatic disease is assumed to be either 0% (A), 25% (B), 50% (C), or 75% (D).

**Supplementary Figure 6:**
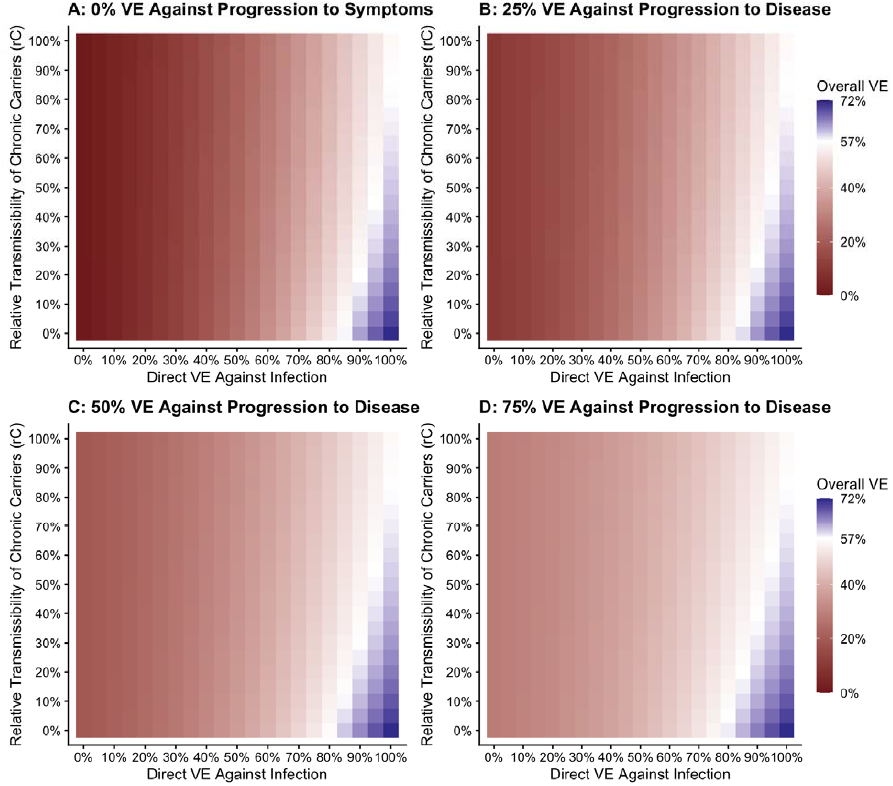
Projected overall trial effect by direct vaccine efficacy against infection and the transmissibility of chronic carriers, alternative model. This figure demonstrates the impact of *VE*_*progression*_ on overall vaccine protection in the alternative model. Each pixel represents the projected overall protection in the TyVAC-Bangladesh trial for a given level of TCV efficacy against *S*. Typhi infection (x-axis) and relative transmissibility of chronic carriers compared to acute cases (y-axis). In each panel, TCV efficacy against progression from infection to symptomatic disease (*VE*_*progression*_) is assumed to be either 0% (A), 25% (B), 50% (C), or 75% (D).

**Supplementary Figure 7:**
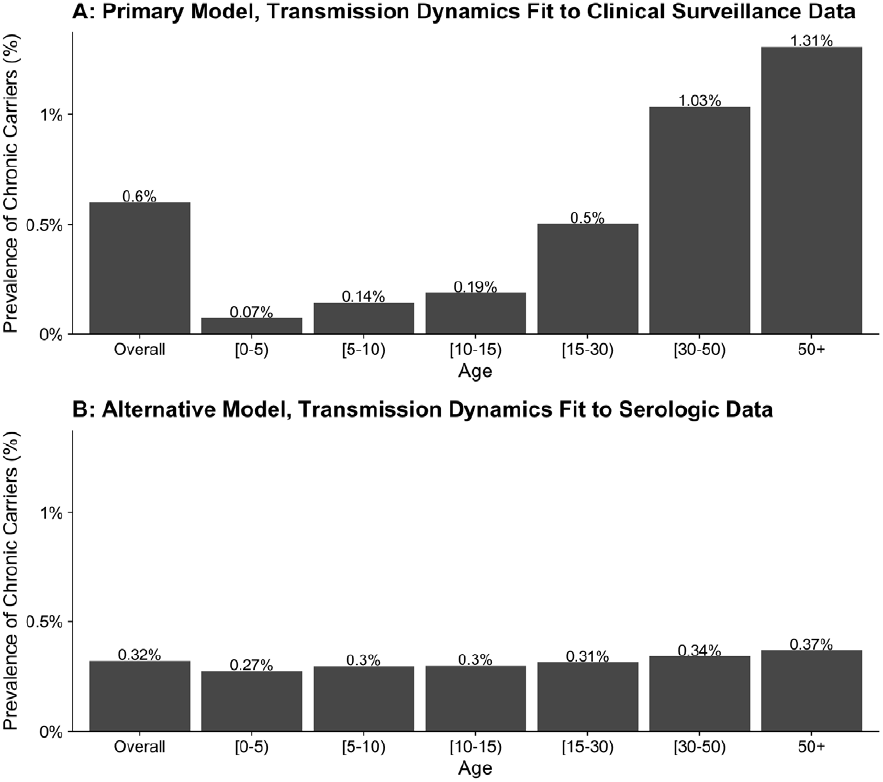
Modeled prevalence of chronic carriers in the absence of vaccination. Bars represent the overall and age-specific percent of the population who are chronic carriers in the primary model (A) and the alternative model (B).

**Supplementary Figure 8:**
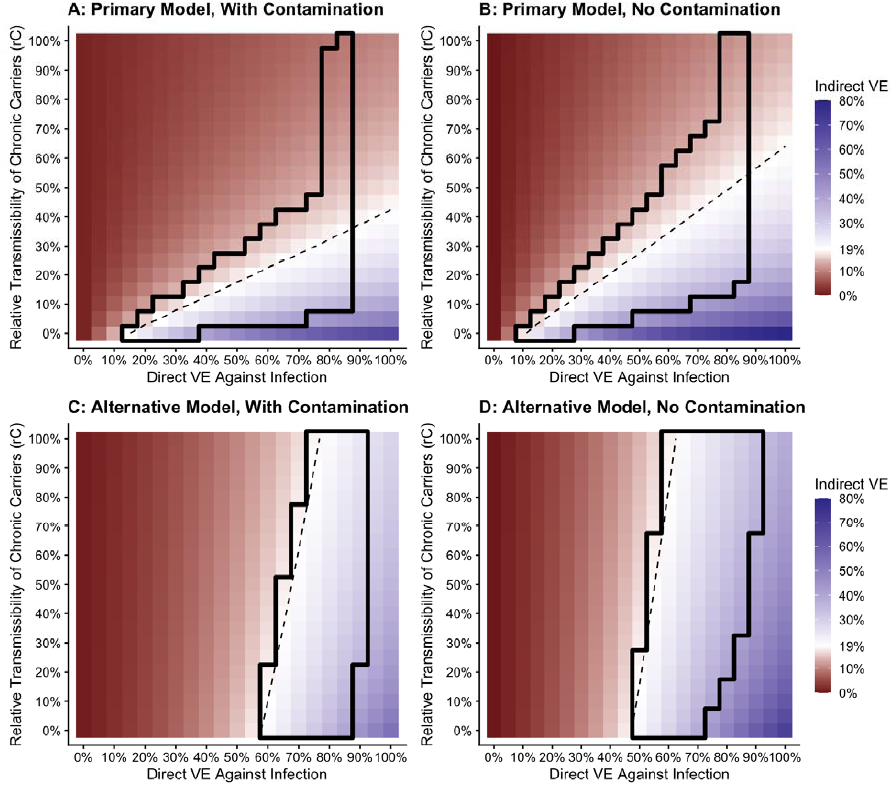
Projected indirect trial effect by direct vaccine efficacy against infection and the transmissibility of chronic carriers, with and without contamination across cluster boundaries. This figure demonstrates the impact of contamination on indirect vaccine protection in the primary and alternative models. Panels in the top (A,B) and bottom (C,D) rows correspond to the primary and alternative model, respectively. Panels in the left column (A, C) display the results of the main analysis, in which there is some contamination (transmission across cluster boundaries), while the panels in the right column (B, D) are based on a sensitivity analysis in which there is no contamination. Each pixel represents the projected indirect protection in the TyVAC-Bangladesh trial for a given level of TCV efficacy against *S*. Typhi infection (x-axis) and relative transmissibility of chronic carriers compared to acute cases (y-axis). The region bounded by the solid black line contains parameter combinations for which model outputs are consistent with the estimated total, overall, and indirect protection in the TyVAC-Bangladesh trial. The dashed line interpolates parameter sets which reproduce the 19% trial estimate of indirect protection.

**Supplementary Figure 9:**
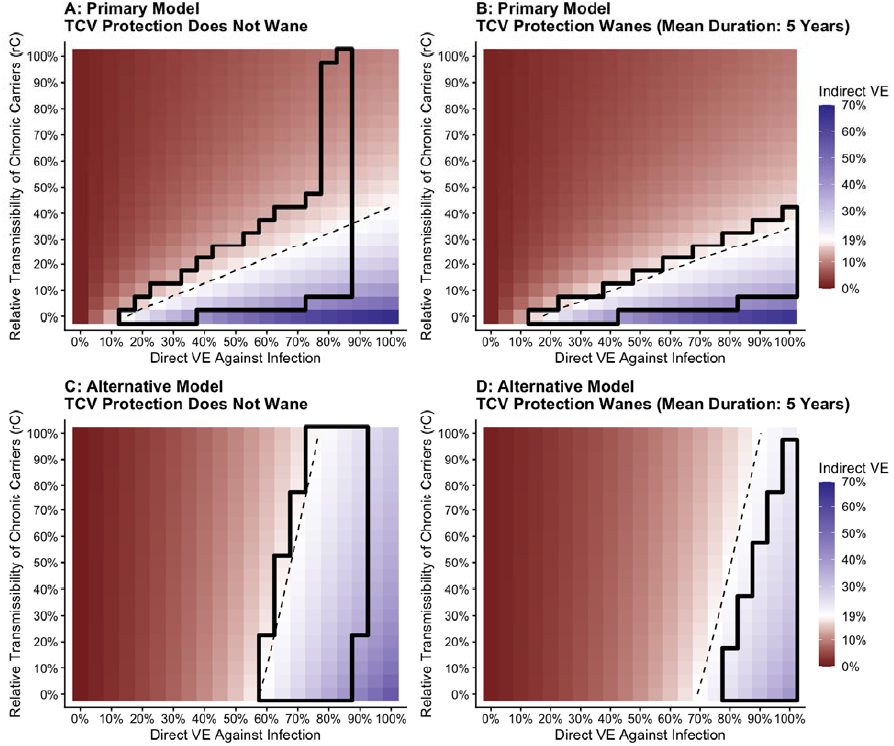
Projected indirect trial effect by direct vaccine efficacy against infection and the transmissibility of chronic carriers, assuming waning vaccine protection over time. This figure demonstrates the impact of waning TCV immunity on indirect vaccine protection in the primary and alternative models. Panels in the top (A,B) and bottom (C,D) rows correspond to the primary and alternative model, respectively. Panels in the left column (A, C) display the results of the main analysis, in which TCV protection does not wane, while the panels in the right column (B, D) are based on a sensitivity analysis in which the duration of vaccine protection is exponentially distributed, with a mean duration of five years. Each pixel represents the projected indirect protection in the TyVAC-Bangladesh trial for a given level of TCV efficacy against *S*. Typhi infection (x-axis) and relative transmissibility of chronic carriers compared to acute cases (y-axis). The region bounded by the solid black line contains parameter combinations for which model outputs are consistent with the estimated total, overall, and indirect protection in the TyVAC-Bangladesh trial. The dashed line interpolates parameter sets which reproduce the 19% trial estimate of indirect protection.

## References

1. Meiring JE, Khanam F, Basnyat B, Charles RC, Crump JA, Debellut F, et al. Typhoid fever. Nat Rev Dis Prim. 2023;9(1):71. Available from: 10.1038/s41572-023-00480-z

2. Antillón M, Warren JL, Crawford FW, Weinberger DM, Kürüm E, Pak GD, et al. The burden of typhoid fever in low- and middle-income countries: A meta-regression approach. PLoS Negl Trop Dis. 2017 Feb 27;11(2):e0005376. Available from: 10.1371/journal.pntd.0005376

3. Batool R, Qamar ZH, Salam RA, Yousafzai MT, Ashorn P, Qamar FN. Efficacy of typhoid vaccines against culture-confirmed S<em>almonella</em> Typhi in typhoid endemic countries: a systematic review and meta-analysis. Lancet Glob Heal. 2024 Apr 1;12(4):e589–98. Available from: 10.1016/S2214-109X(23)00606-X

4. Hancuh M, Walldorf J, Minta AA, Tevi-Benissan C, Christian KA, Nedelec Y, et al. Typhoid Fever Surveillance, Incidence Estimates, and Progress Toward Typhoid Conjugate Vaccine Introduction — Worldwide, 2018–2022. Morb Mortal Wkly Rep. 2023;72(7):171–6.

5. Debellut F, Bello G, Chisema M, Mkisi R, Kamzati M, Pecenka C, et al. Cost of the typhoid conjugate vaccine introduction through an integrated campaign and follow-on routine immunization in Malawi. Vaccine X. 2024;21:100583. Available from: https://www.sciencedirect.com/science/article/pii/S2590136224001566

6. Neya CO. Reaching 10 million children in Burkina Faso with TCVs. Take On Typhoid Blog. 2025. Available from: https://www.coalitionagainsttyphoid.org/reaching-10-million-children-in-burkina-faso-with-tcvs/

7. 16 million children protected in Kenya’s measles, rubella and typhoid vaccination campaign. World Health Organization - Regional Office for Africa. 2025. Available from: https://www.afro.who.int/node/22070

8. Bangladesh launches nationwide typhoid conjugate vaccine campaign to protect 50 million children. Gavi - The Vaccine Alliance. 2025. Available from: https://www.gavi.org/news/media-room/bangladesh-launches-nationwide-typhoid-conjugate-vaccine-campaign-protect-50

9. Burrows H, Antillón M, Gauld JS, Kim J-H, Mogasale V, Ryckman T, et al. Comparison of model predictions of typhoid conjugate vaccine public health impact and cost-effectiveness. Vaccine. 2023;41(4):965–75. Available from: https://www.sciencedirect.com/science/article/pii/S0264410X22015572

10. Dipika S, Leon OR, K. BS, K. GN, Mohammad A, Byomkesh M, et al. A Cluster-Randomized Effectiveness Trial of Vi Typhoid Vaccine in India. N Engl J Med. 2009 May 12;361(4):335– 44. Available from: 10.1056/NEJMoa0807521

11. Khan MI, Soofi SB, Ochiai RL, Habib MA, Sahito SM, Nizami SQ, et al. Effectiveness of Vi capsular polysaccharide typhoid vaccine among children: A cluster randomized trial in Karachi, Pakistan. Vaccine. 2012;30(36):5389–95. Available from: https://www.sciencedirect.com/science/article/pii/S0264410X12008560

12. Theiss-Nyland K, Shakya M, Colin-Jones R, Voysey M, Smith N, Karkey A, et al. Assessing the Impact of a Vi-polysaccharide Conjugate Vaccine in Preventing Typhoid Infections Among Nepalese Children: A Protocol for a Phase III, Randomized Control Trial. Clin Infect Dis. 2019 Mar 7;68(Supplement_2):S67–73. Available from: 10.1093/cid/ciy1106

13. Shiri T, Datta S, Madan J, Tsertsvadze A, Royle P, Keeling MJ, et al. Indirect effects of childhood pneumococcal conjugate vaccination on invasive pneumococcal disease: a systematic review and meta-analysis. Lancet Glob Heal. 2017 Jan 1;5(1):e51–9. Available from: 10.1016/S2214-109X(16)30306-0

14. Barbour ML, Mayon-White RT, Coles C, Crook DW, Moxon ER. The impact of conjugate vaccine on carriage of Haemophilus influenzae type b. J Infect Dis. 1995 Jan;171(1):93–8.

15. Kristiansen PA, Diomandé F, Ba AK, Sanou I, Ouédraogo A, Ouédraogo R, et al. Impact of the Serogroup A Meningococcal Conjugate Vaccine, MenAfriVac, on Carriage and Herd Immunity. Clin Infect Dis. 2012 Oct 19;56(3):354–63. Available from: 10.1093/cid/cis892

16. Voysey M, Pollard AJ. Seroefficacy of Vi Polysaccharide–Tetanus Toxoid Typhoid Conjugate Vaccine (Typbar TCV). Clin Infect Dis. 2018 Jun 18;67(1):18–24. Available from: 10.1093/cid/cix1145

17. Gibani MM, Voysey M, Jin C, Jones C, Thomaides-Brears H, Jones E, et al. The Impact of Vaccination and Prior Exposure on Stool Shedding of Salmonella Typhi and Salmonella Paratyphi in 6 Controlled Human Infection Studies. Clin Infect Dis. 2019 Apr 8;68(8):1265–73. Available from: 10.1093/cid/ciy670

18. Qadri F, Khanam F, Liu X, Theiss-Nyland K, Biswas PK, Bhuiyan AI, et al. Protection by vaccination of children against typhoid fever with a Vi-tetanus toxoid conjugate vaccine in urban Bangladesh: a cluster-randomised trial. Lancet. 2021 Aug 21;398(10301):675– 84. Available from: 10.1016/S0140-6736(21)01124-7

19. Khanam F, Kim DR, Liu X, Voysey M, Pitzer VE, Zaman K, et al. Assessment of vaccine herd protection in a cluster-randomised trial of Vi conjugate vaccine against typhoid fever: results of further analysis. eClinicalMedicine. 2023 Apr 1;58. Available from: 10.1016/j.eclinm.2023.101925

20. Walker J, Russell P, Kermack L, Van TT, Nga TVT, Mylona E, et al. Leveraging paired serology to estimate the incidence of typhoidal <em>Salmonella</em> infection in the STRATAA study. medRxiv. 2025 Jan 1;2025.03.15.25324021. Available from: http://medrxiv.org/content/early/2025/03/17/2025.03.15.25324021.abstract

21. Pitzer VE, Bowles CC, Baker S, Kang G, Balaji V, Farrar JJ, et al. Predicting the Impact of Vaccination on the Transmission Dynamics of Typhoid in South Asia: A Mathematical Modeling Study. PLoS Negl Trop Dis. 2014 Jan 9;8(1):e2642. Available from: 10.1371/journal.pntd.0002642

22. Meiring JE, Shakya M, Khanam F, Voysey M, Phillips MT, Tonks S, et al. Burden of enteric fever at three urban sites in Africa and Asia: a multicentre population-based study. Lancet Glob Heal. 2021 Dec 1;9(12):e1688–96. Available from: 10.1016/S2214-109X(21)00370-3

23. Mylona E, Hefele L, Tran Vu Thieu N, Trinh Van T, Nguyen Ngoc Minh C, Tran Tuan A, et al. The Identification of Enteric Fever-Specific Antigens for Population-Based Serosurveillance. J Infect Dis. 2024 Mar 15;229(3):833–844. Available from: 10.1093/infdis/jiad242

24. Bilcke J, Antillón M, Pieters Z, Kuylen E, Abboud L, Neuzil KM, et al. Cost-effectiveness of routine and campaign use of typhoid Vi-conjugate vaccine in Gavi-eligible countries: a modelling study. Lancet Infect Dis. 2019 Jul 1;19(7):728–39. Available from: 10.1016/S1473-3099(18)30804-1

25. Kaufhold S, Yaesoubi R, Pitzer VE. Predicting the Impact of Typhoid Conjugate Vaccines on Antimicrobial Resistance. Clin Infect Dis. 2019 Mar 7;68(Supplement_2):S96–104. Available from: 10.1093/cid/ciy1108

26. Pitzer VE, Feasey NA, Msefula C, Mallewa J, Kennedy N, Dube Q, et al. Mathematical Modeling to Assess the Drivers of the Recent Emergence of Typhoid Fever in Blantyre, Malawi. Clin Infect Dis. 2015 Nov 1;61(Suppl_4):S251–8. Available from: 10.1093/cid/civ710

27. Saad NJ, Bowles CC, Grenfell BT, Basnyat B, Arjyal A, Dongol S, et al. The impact of migration and antimicrobial resistance on the transmission dynamics of typhoid fever in Kathmandu, Nepal: A mathematical modelling study. PLoS Negl Trop Dis. 2017 May 5;11(5):e0005547. Available from: 10.1371/journal.pntd.0005547

28. B. HR, E. GS, E. WT, L. DH, T. DA, J. SM. Typhoid Fever: Pathogenesis and Immunologic Control. N Engl J Med. 1970 Sep 24;283(13):686–91. Available from: 10.1056/NEJM197009242831306

29. Ames WR, Robins M. Age and Sex as Factors in the Development of the Typhoid Carrier State, and a Method for Estimating Carrier Prevalence. Am J Public Heal Nations Heal. 1943 Mar 1;33(3):221–30. Available from: 10.2105/AJPH.33.3.221

30. Halloran ME, Longini Jr. IM, Struchiner CJ. Design and Interpretation of Vaccine Field Studies. Epidemiol Rev. 1999 Jan 1;21(1):73–88. Available from: 10.1093/oxfordjournals.epirev.a017990

31. Shakya M, Voysey M, Theiss-Nyland K, Colin-Jones R, Pant D, Adhikari A, et al. Efficacy of typhoid conjugate vaccine in Nepal: final results of a phase 3, randomised, controlled trial. Lancet Glob Heal. 2021 Nov 1;9(11):e1561–8. Available from: 10.1016/S2214-109X(21)00346-6

32. Patel PD, Liang Y, Meiring JE, Chasweka N, Patel P, Misiri T, et al. Efficacy of typhoid conjugate vaccine: final analysis of a 4-year, phase 3, randomised controlled trial in Malawian children. Lancet. 2024 Feb 3;403(10425):459–68. Available from: 10.1016/S0140-6736(23)02031-7

33. Jin C, Gibani MM, Moore M, Juel HB, Jones E, Meiring J, et al. Efficacy and immunogenicity of a Vi-tetanus toxoid conjugate vaccine in the prevention of typhoid fever using a controlled human infection model of Salmonella Typhi: a randomised controlled, phase 2b trial. Lancet. 2017;390(10111):2472–80. Available from: https://www.sciencedirect.com/science/article/pii/S0140673617321499

34. Yousafzai MT, Karim S, Qureshi S, Kazi M, Memon H, Junejo A, et al. Effectiveness of typhoid conjugate vaccine against culture-confirmed <em>Salmonella enterica</em> serotype Typhi in an extensively drug-resistant outbreak setting of Hyderabad, Pakistan: a cohort study. Lancet Glob Heal. 2021 Aug 1;9(8):e1154–62. Available from: 10.1016/S2214-109X(21)00255-2

35. Lightowler MS, Manangazira P, Nackers F, Van Herp M, Phiri I, Kuwenyi K, et al. Effectiveness of typhoid conjugate vaccine in Zimbabwe used in response to an outbreak among children and young adults: A matched case control study. Vaccine. 2022;40(31):4199–210. Available from: https://www.sciencedirect.com/science/article/pii/S0264410X22005527

36. Hoffman SA, LeBoa C, Date K, Haldar P, Harvey P, Shimpi R, et al. Programmatic Effectiveness of a Pediatric Typhoid Conjugate Vaccine Campaign in Navi Mumbai, India. Clin Infect Dis. 2023 Jul 1;77(1):138–44. Available from: 10.1093/cid/ciad132

37. World Health Organization. Typhoid vaccines: WHO position paper - March 2018. Wkly Epidemiol Rec. 2018;93(13).

38. GAVI Zero-Dose Learning Hub. Introduction of Typhoid Conjugate Vaccine (TCV) in Nepal: Role of the Vaccination Campaign in Identifying and Reaching Zero-Dose Children. 2023.

39. Qadri F, Khanam F, Zhang Y, Biswas PK, Voysey M, Mujadidi YF, et al. 5-year vaccine protection following a single dose of Vi-tetanus toxoid conjugate vaccine in Bangladeshi children (TyVOID): a cluster randomised trial. Lancet. 2024 Oct 12;404(10461):1419–29. Available from: 10.1016/S0140-6736(24)01494-6

40. Khanam F, Darton TC, Meiring JE, Kumer Sarker P, Kumar Biswas P, Bhuiyan MAI, et al. Salmonella Typhi Stool Shedding by Patients With Enteric Fever and Asymptomatic Chronic Carriers in an Endemic Urban Setting. J Infect Dis. 2021 Dec 15;224(Supplement_7):S759–63. Available from: 10.1093/infdis/jiab476

41. Charles RC, Sultana T, Alam MM, Yu Y, Wu-Freeman Y, Bufano MK, et al. Identification of Immunogenic Salmonella enterica Serotype Typhi Antigens Expressed in Chronic Biliary Carriers of S. Typhi in Kathmandu, Nepal. PLoS Negl Trop Dis. 2013 Aug 1;7(8):e2335. Available from: 10.1371/journal.pntd.0002335

42. Nolan C, White P, Feeley J, Brown S, Hambie E, Wong K-H. Vi Serology in the Detection of Typhoid Carriers. Lancet. 1981;317(8220):583–5. Available from: https://www.sciencedirect.com/science/article/pii/S014067368192033X

43. Birger R, Antillón M, Bilcke J, Dolecek C, Dougan G, Pollard AJ, et al. Estimating the effect of vaccination on antimicrobial-resistant typhoid fever in 73 countries supported by Gavi: a mathematical modelling study. Lancet Infect Dis. 2022 May 1;22(5):679–91. Available from: 10.1016/S1473-3099(21)00627-7

44. Feng S, Zhang Y, Khanam F, Voysey M, Pitzer VE, Qadri F, et al. The validity of test-negative design for assessment of typhoid conjugate vaccine protection: comparison of estimates by different study designs using data from a cluster-randomised controlled trial. Lancet Glob Heal. 2025 Jun;13(6):e1122–31.

